# A Retrospective Analysis of COVID-19 mRNA Vaccine Breakthrough Infections – Risk Factors and Vaccine Effectiveness

**DOI:** 10.1101/2021.10.05.21264583

**Authors:** Cong Liu, Junghwan Lee, Casey Ta, Ali Soroush, James R. Rogers, Jae Hyun Kim, Karthik Natarajan, Jason Zucker, Chunhua Weng

**Author notes:** Correspondence Chunhua Weng, PhD, Department of Biomedical Informatics, Columbia University, 622 W 168 Street, PH-20, Room 407, New York, NY 10032.

## Abstract

**Importance:** Little is known about COVID vaccine breakthrough infections and their risk factors.

**Objective:** To identify risk factors associated with COVID-19 breakthrough infections among vaccinated individuals and to reassess the effectiveness of COVID-19 vaccination against severe outcomes using real-world data.

**Design, Setting, and Participants:** We conducted a series of observational retrospective analyses using the electronic health records (EHRs) of Columbia University Irving Medical Center/New York Presbyterian (CUIMC/NYP) up to September 21, 2021. New York adult residence with PCR test records were included in this analysis.

**Main Outcomes and Measures:** Poisson regression was used to assess the association between breakthrough infection rate in vaccinated individuals and multiple risk factors – including vaccine brand, demographics, and underlying conditions – while adjusting for calendar month, prior number of visits and observational days. Logistic regression was used to assess the association between vaccine administration and infection rate by comparing a vaccinated cohort to a historically matched cohort in the pre-vaccinated period. Infection incident rate was also compared between vaccinated individuals and longitudinally matched unvaccinated individuals. Cox regression was used to estimate the association of the vaccine and COVID-19 associated severe outcomes by comparing breakthrough cohort and two matched unvaccinated infection cohorts.

**Results:** Individuals vaccinated with Pfizer/BNT162b2 (IRR against Moderna/mRNA-1273 [95% CI]: 1.66 [1.17 – 2.35]); were male (1.47 [1.11 – 1.94%]); and had compromised immune systems (1.48 [1.09 – 2.00]) were at the highest risk for breakthrough infections. Vaccinated individuals had a significant lower infection rate among all subgroups. An increased incidence rate was found in both vaccines over the time. Among individuals infected with COVID-19, vaccination significantly reduced the risk of death (adj. HR: 0.20 [0.08 - 0.49]).

**Conclusion and Relevance:** While we found both mRNA vaccines were effective, Moderna/mRNA-1273 had a lower incidence rate of breakthrough infections. Both vaccines had increased incidence rates over the time. Immunocompromised individuals were among the highest risk groups experiencing breakthrough infections. Given the rapidly changing nature of the SARS-CoV-2, continued monitoring and a generalizable analysis pipeline are warranted to inform quick updates on vaccine effectiveness in real time.

**Key Points:** *Question:* What risk factors contribute to COVID-19 breakthrough infections among mRNA vaccinated individuals? How do clinical outcomes differ between vaccinated but still SARS-CoV-2 infected individuals and non-vaccinated, infected individuals?

*Findings:* This retrospective study uses CUIMC/NYP EHR data up to September 21, 2021. Individuals who were vaccinated with Pfizer/BNT162b2, male, and had compromised immune systems had significantly higher incidence rate ratios of breakthrough infections. Comparing demographically matched pre-vaccinated and unvaccinated individuals, vaccinated individuals had a lower incidence rate of SARS-CoV-2 infection among all subgroups.

*Meaning:* Leveraging real-world EHR data provides insight on who may optimally benefit from a booster COVID-19 vaccination.

## INTRODUCTION

The ongoing global COVID-19 pandemic has infected hundreds of millions of people over the world, imposing a tremendous burden on the global healthcare system. COVID-19 vaccines are currently the best defense against the rapidly evolving severe acute respiratory syndrome coronavirus 2 (SARS-CoV-2), having demonstrated efficacy in preventing symptomatic COVID-19 while being relatively safe in trial studies^1-3^. As of August 2021, about 50% of the total US population had been fully vaccinated^4^. The Centers for Disease Control and Prevention (CDC) have reported a small percentage of fully vaccinated people experiencing vaccine breakthrough infections^5^. However, there are emerging concerns about vaccine breakthrough infections^6^. Studies have been conducted to confirm vaccine breakthrough infections with SARS-CoV-2 variants using genome sequencing^7^ and to investigate clinical characteristics of the vaccine breakthrough infections^8-10^. Here, we retrospectively analyzed electronic health records (EHRs) from Columbia University Irving Medical Center/New York-Presbyterian (CUIMC/NYP), which is part of the CDC-organized VISION Network for COVID-19 vaccine effectiveness assessment^11^. We aimed to address the following three questions: (1) what risk factors are associated with breakthrough infection; (2) how effective are vaccines in reducing the rate of SARS-CoV-2 infection in our available population; and (3) how effective are vaccines in reducing the risk of SARS-CoV-2 associated severe outcomes among the infected population.

## METHODS

### Study design and population

We used EHR data obtained from the NYP/CUIMC data warehouse. NYP/CUIMC is a quaternary care academic medical center that includes an academic hospital, children’s hospital, and community-based hospital serving a diverse patient population in northern Manhattan, New York City (NYC). EHR data were collected and stored in the data warehouse during routine clinical care at CUIMC/NYP. The EHR data is converted to the Observational Medical Outcomes Partnership (OMOP) Common Data Model (CDM)^12^. All data involved in this analysis were collected up to September 21, 2021.

### Cohort Definition

Only individuals older than 18 years residing in NYC were included for this study. OMOP concepts related to vaccines were used to identify vaccinated individuals who received two doses of Pfizer/BNT162b2 or Moderna/mRNA-1273. To minimize potential bias resulting from missing vaccination records, vaccines records in our data warehouse were obtained from both CUIMC EHR data and the NYC vaccine registry. The two-dose interval requirement is 20-23 days for Pfizer/BNT162b2, and 27-31 days for Moderna/mRNA-1273; individuals with two doses with 14 days of available follow-up after their second dose were considered fully vaccinated. Individuals who received doses from more than one manufacturer or only received one vaccine dose were excluded. OMOP measurement concepts and corresponding value concepts related to detect positive RNA were used to identify individuals with at least one SARS-CoV-2 PCR test. Individuals with a positive SARS-CoV-2 PCR test, a positive SARS-CoV-2 antibody test, or a concept indicating a SARS-CoV-2 infection in the condition table were flagged as having evidence of SARS-CoV-2 infection. The details of OMOP concepts used for cohort definition is available in **eFile 1**.

Based on the vaccine and SARS-CoV-2 status, we then constructed six cohorts as shown in **Figure 1** (with **eFigure 1-3** providing more detailed breakdowns for each cohort); in brief, they are as follows:

**Figure 1.**
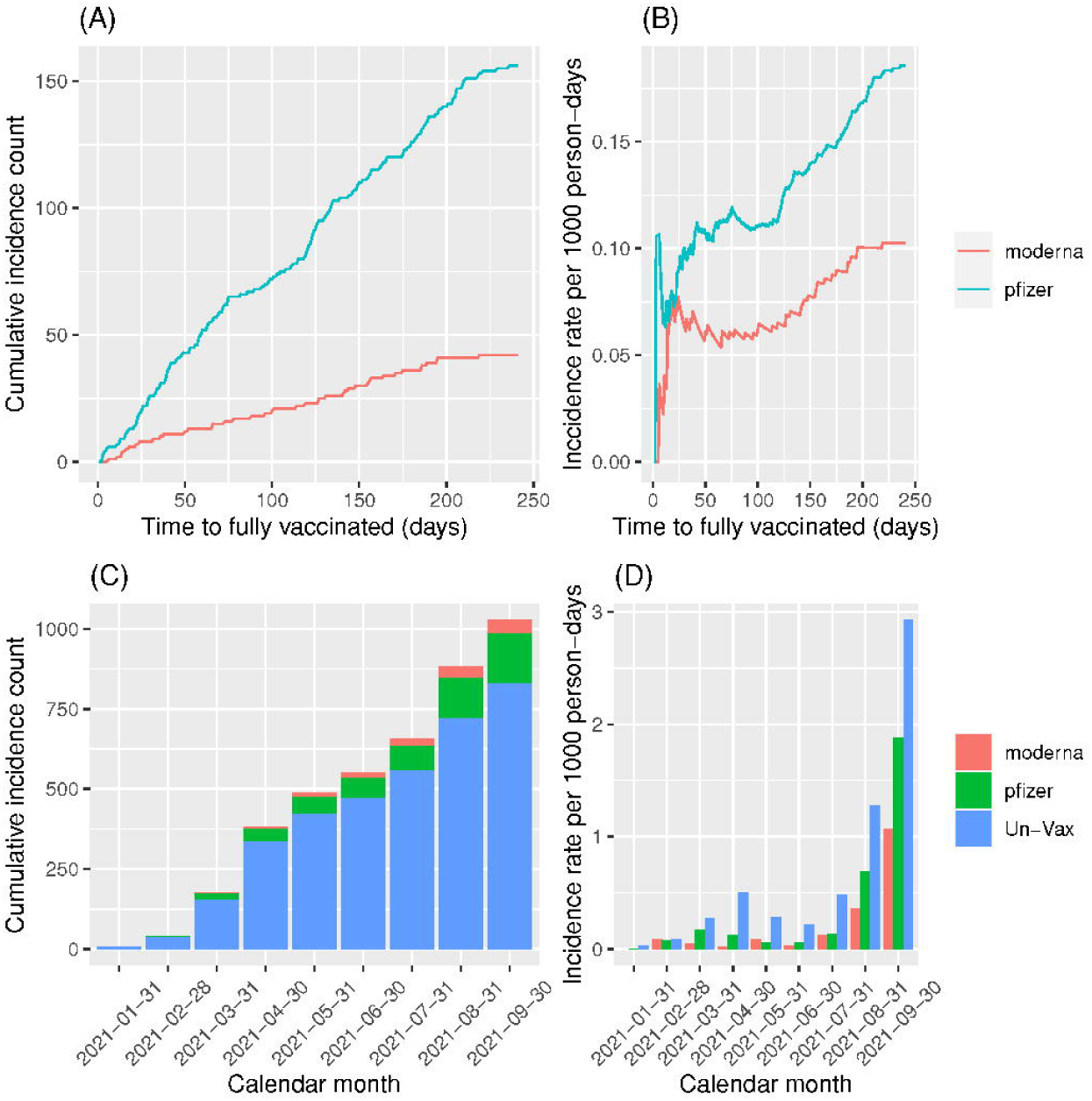
Cohort construction diagram and study overview. Vaccines records were obtained both from CUIMC EHR data and the NYC vaccine registry. Only fully vaccinated individuals with mRNA vaccines were included. Individuals Individual with positive SARS-CoV-PCR test, a positive SARS-CoV-2 antibody test, or a concept indicating a SARS-CoV-2 infection in the condition table were flagged as having evidences of SARS-CoV-2 infection. No COVID evidences were required before entering the cohort for positive individuals and before exiting the cohort for negative individuals. Only age > 18 and NYC residents were included in this analysis. **“**vax”: Individuals 14 days after receiving their second doses who considered as fully vaccinated; “EUA”: The date first dose of vaccine is administrated (i.e. Dec 11 2021); “1^st^ Dose”: the date of the individual was administrated his/her first (including J&J) vaccine dose (or end of study if vaccine was not ever administrated).

- “Vax” Cohorts: These two mutually exclusive cohorts refer to vaccinated individuals. The entry date is January 18, 2020 (the first individual fully vaccinated). End date is Sep 21, 2021.
  - “Vax positive” (N = 198): Individuals with a positive PCR test after full vaccination and without evidence of SARS-CoV-2 infection before full vaccination.
  - “Vax negative” (N = 14,164): Individuals with a negative PCR test after full vaccination and without evidence of SARS-CoV-2 infection at any time in their records.
- “Pre-Vax” Cohorts: These two mutually exclusive cohorts refer to individuals at risk of infection with SARS-CoV-2 during a time period when a vaccine was unavailable. The first allowed entry is January 1, 2020 while the calendar end date is December 10, 2020 (first dose available). Individuals in a “Pre-Vax” negative cohort can also be in a “Vax” cohort.
  - “Pre-Vax positive” (N = 6,462): Individuals with a positive PCR test before the vaccination period.
  - “Pre-Vax negative” (N = 55,580): Individuals with a negative PCR test and without any evidence of SARS-CoV-2 infection before the vaccination period.
- “Un-Vax” Cohorts: These two mutually exclusive cohorts refer to individuals at risk of infection with SARS-CoV-2 during the time period when vaccines were available, but did not have a vaccine administered. The entry date is the same as the “Vax” cohorts. Of note, if anindividual receives a first dose for vaccination, that individual exits the “Un-Vax” cohort (and may later become part of a “Vax” cohort).
  - “Un-Vax positive” (N = 3,902): Individuals with a positive PCR test after entry date and before administration of a first vaccination dose (if ever administrated), while having no evidence of SARS-CoV-2 infection before entry date.
  - “Un-Vax negative” (N = 33,850): Individuals with a negative PCR test after entry date and before administration of a first vaccination dose (if ever administrated), while having no evidence of SARS-CoV-2 infection before entry date.

### Feature Extraction

For each cohort, we extracted individuals’ demographic data including age, sex, ethnicity, and race. For the vaccinated cohorts, vaccine brand and corresponding administration dates were also extracted. To approximate available observation time, we extracted the total number of prior EHR visits, and days of observation periods between clinical encounters for each individual. We extracted all previous condition and drug concepts from the *condition_era* and *drug_era* tables. To avoid extracting condition/drug concepts potentially caused by the SARS-CoV-2 infection itself, we added a 90-day washout period (i.e. ignore all the concepts within the 90-day window prior to the PCR test regardless of its result). To identify individuals who might have compromised immune systems, we compiled a list of conditions and drugs including active solid tumor and hematologic malignancies (within two years), solid-organ or hematopoietic stem cell transplant, primary immunodeficiencies, human immunodeficiency virus (HIV) infection, immunosuppressive therapies (e.g., cancer chemotherapeutic agents, certain biologic agents, rituximab, etc.) and chronic kidney disease (CKD)^13^ (**eTable 1**); individuals can fall into multiple immunocompromised subgroups. To adjust for the caseload in NYC, 7-day rolling average of cases was applied^14^. We extracted the clinical outcomes of interest, including COVID-19 associated hospitalization (defined as within 28 days after, or 72 hours before getting a positive PCR result), mechanical ventilation, tracheostomy, and death as severe outcomes of the infection based on the World Health Organization’s clinical progression scale for COVID-19^15^. We did not extract intensive care unit (ICU) admissions because many patients were admitted into repurposed units for ICU-level care during the pandemic, leading to missing ICU reporting in the EHR.

### Identifying risk factors associated with breakthrough infection

We compared the “Vax positive” and “Vax negative” cohorts to identify potential risk factors associated with breakthrough infections (**eFigure 1**). The entry date was defined as fully vaccinated date, and individuals were then followed until the first positive PCR date (or end of the study for “Vax negative” individuals). For each risk factor (e.g., vaccine brand, demographics, immunocompromised status), a univariate Poisson regression was fit to assess the incidence rate ratio (IRR; i.e., breakthrough per 1,000 person-days) against the reference status. To minimize potential bias resulting from daily caseload, viral mutations, and EHR data quality, the Poisson regression was adjusted for: (1) total number of observation days in the EHR before the entry date; (2) total number of visits in the EHR before the entry date; and (3) calendar month of the PCR test date. We further applied a non-hypothesis-driven approach to uniformly evaluate the risk effect for each historical condition and drug by fitting a univariate Poisson regression with similar adjustment. Condition and drug concepts significantly associated with the breakthrough infections were identified as < 0.05 Bonferroni adjusted p-value^16^.

### Evaluate vaccine effectiveness in preventing infection by comparing vaccinated individuals with pre- and un-vaccinated individuals

For the “Vax” cohorts, the entry date was defined similarly to the entry date for the risk factor analysis. For the “Un-Vax” cohorts, the entry date was defined as Jan 18^th^, 2021 (14 days after the first individual received their second dose at CUIMC/NYP), and individuals were then followed up until the first positive PCR test (latest for negative individuals) or the date when they received their first dose, whichever came first (**eFigure 2**). We 1:1 matched vaccinated individuals to unvaccinated individuals using a nearest neighbor search based on (1) observation days; (2) visit count; (3) calendar week of the PCR test (earliest positive PCR or latest negative PCR); (4) demographics (e.g., sex, age, race, ethnicity); and (5) immunocompromised status (binary). The IRRs for the vaccine were estimated via Poisson regressions.

We further identified 1:1 matched individuals in the “Pre-Vax” cohort based on the same covariates, except for the calendar week of the PCR test, which was replaced by the 7-day rolling average of cases in NYC at the PCR testing date. Given the difficulty in identifying an appropriate entry date for the pre-vaccinated cohort, we applied a case/control design to calculate the odds ratio (OR) of contracting COVID-19 infection between “Vax” cohort and “Pre-Vax” cohort using logistic regressions.

### Evaluate vaccine effectiveness in preventing severe outcomes among infected individuals

We compared hazard ratios (HRs) of experiencing severe clinical outcomes between the “Vax positive” and matched “Un-Vax positive”/”Pre-Vax positive” cohort (1:10 ratio for each). The severe outcomes were defined as SARS-CoV-2 associated severe clinical events, including hospitalization (includes emergency room visits), mechanical ventilation, tracheostomy, and death (**eTable 1**). The entry date for each cohort was defined as the earliest qualified PCR positive test date, and follow-up time ended at either outcome, the last date of the patient’s visiting records in our medical system, end of study, or 28 days after the PCR positive results, whichever came first. HRs for vaccine effectiveness were estimated via Cox regressions.

## RESULTS

Table 1 provides baseline characteristics of the six cohorts (note: some individuals are in multiple cohorts at different times). For the “Vax positive” (i.e., breakthrough) cohort, the average age was 58.5 (SD: 20.3). 156 (78.8%) received Pfizer/BNT162b2, while 42 (21.2%) received Moderna/mRNA-1273. 65 (45.5%) had underlying immunocompromised conditions. 120 (60.6%) of the patients with breakthrough infections were hospitalized. In general, PCR positive individuals had a higher number of prior visits and observational days compared to unvaccinated individuals. For later analyses, we used a matching strategy to balance the covariates.

**Table 1.** Baseline characteristics of the individuals in six cohorts (pre-matched).

|  | “Vax” cohort |  | “Un-Vax” cohort |  | “Pre-Vax” cohort |  |
| --- | --- | --- | --- | --- | --- | --- |
|  | Vax positive | Vax negative | Un-Vax positive | Un-Vax negative | Pre-Vax positive | Pre-Vax negative |
| **N** | 198 | 14164 | 3902 | 33850 | 6462 | 55580 |
|  | Either mean (SD) or N (%) |  |  |  |  |  |
| **Previous encounters** | 80 (124.75) | 65.7 (121.91) | 64 (121.15) | 44.4 (91.4) | 70.6 (127.86) | 45.2 (95.1) |
| **Observational days** | 5470 (3909.61) | 5425.2 (3843.99) | 5940.6 (4045.09) | 4999.3 (3799.28) | 5942.1 (3978.71) | 4932.6 (3672.63) |
| **Age** | 58.5 (20.34) | 59.4 (18.86) | 54.2 (20.06) | 50.9 (19.73) | 58.9 (19.46) | 52.2 (19.79) |
| **Sex** |  |  |  |  |  |  |
| Female | 110(55.6%) | 9010(63.6%) | 2199(56.4%) | 21065(62.2%) | 3293(51%) | 34563(62.2%) |
| Male | 88(44.4%) | 5153(36.4%) | 1702(43.6%) | 12765(37.7%) | 3168(49%) | 21009(37.8%) |
| Unknown/Other | 0 (0%) | 1(0%) | 1(0%) | 20(0.1%) | 1(0%) | 8(0%) |
| **Race** |  |  |  |  |  |  |
| Asian | 3(1.5%) | 545(3.8%) | 73(1.9%) | 804(2.4%) | 132(2%) | 2021(3.6%) |
| Black | 30(15.2%) | 1851(13.1%) | 831(21.3%) | 7046(20.8%) | 1231(19%) | 9218(16.6%) |
| White | 88(44.4%) | 6325(44.7%) | 887(22.7%) | 9740(28.8%) | 1779(27.5%) | 20816(37.5%) |
| Unknown/Other | 77(38.9%) | 5443(38.4%) | 2111(54.1%) | 16260(48%) | 3320(51.4%) | 23525(42.3%) |
| **Ethnicity** |  |  |  |  |  |  |
| Hispanic or Latino | 58(29.3%) | 3932(27.8%) | 1840(47.2%) | 12081(35.7%) | 2823(43.7%) | 15018(27%) |
| Not Hispanic or Latino | 101(51%) | 7571(53.5%) | 1339(34.3%) | 14512(42.9%) | 2224(34.4%) | 27194(48.9%) |
| Unknown/Other | 39(19.7%) | 2661(18.8%) | 723(18.5%) | 7257(21.4%) | 1415(21.9%) | 13368(24.1%) |
| **Vaccine Brand** |  |  |  |  |  |  |
| Moderna/mRNA-1273 | 42(21.2%) | 4626(32.7%) | N/A | N/A | N/A | N/A |
| Pfizer/BNT162b2 | 156(78.8%) | 9538(67.3%) | N/A | N/A | N/A | N/A |
| **immunocompromised<sup>1</sup>** |  |  |  |  |  |  |
| Solid tumor | 46(23.2%) | 2354(16.6%) | 274(7%) | 2826(8.3%) | 629(9.7%) | 6702(12.1%) |
| Chronic Kidney Disease | 28(14.1%) | 1486(10.5%) | 364(9.3%) | 2124(6.3%) | 910(14.1%) | 4098(7.4%) |
| HIV | 9(4.5%) | 478(3.4%) | 114(2.9%) | 982(2.9%) | 190(2.9%) | 1603(2.9%) |
| On immunosuppressive therapy | 13(6.6%) | 362(2.6%) | 74(1.9%) | 616(1.8%) | 156(2.4%) | 1248(2.2%) |
| immunodeficiency disorders | 49(24.7%) | 2545(18%) | 370(9.5%) | 3124(9.2%) | 759(11.7%) | 6660(12%) |
| Organ transplant | 10(5.1%) | 366(2.6%) | 108(2.8%) | 610(1.8%) | 244(3.8%) | 1288(2.3%) |
| None | 108(54.5%) | 9031(63.8%) | 3072(78.7%) | 26835(79.3%) | 4641(71.8%) | 41150(74%) |
| **Severe outcomes<sup>1</sup>** |  |  |  |  |  |  |
| Death | 5(2.5%) | 157(1.1%) | 170(4.4%) | 455(1.3%) | 715(11.1%) | 600(1.1%) |
| Tracheostomy | 0 (0%) | 5(0%) | 16(0.4%) | 22(0.1%) | 69(1.1%) | 31(0.1%) |
| Ventilation | 9(4.5%) | 130(0.9%) | 249(6.4%) | 394(1.2%) | 560(8.7%) | 502(0.9%) |
| Hospitalization | 120(60.6%) | 6260(44.2%) | 3031(77.7%) | 18336(54.2%) | 4010(62.1%) | 19579(35.2%) |
| None | 77(38.9%) | 7858(55.5%) | 865(22.2%) | 15422(45.6%) | 2317(35.9%) | 35847(64.5%) |

<sup>1</sup>these are not mutually exclusive (except for the “None” category)

The overall estimated breakthrough infection rate is 0.16 (95% CI: 0.14 – 0.18). **Table 2** summarizes risk factors associated with breakthrough infections. We found a significantly higher incidence rate in vaccinated males than females (IRR = 1.47; 95% CI: 1.11-1.94). We did not find any significant change in incidence rate associated with other demographics, though older and non-Asian individuals are likely to have a higher incidence rate. There was a significant increase in incidence rate among those vaccinated with Pfizer/BNT162b2 compared to Moderna/mRNA-1273 (adj. IRR = 1.66; 95% CI: 1.17-2.35). Underlying compromised immune system were significantly associated with high incident rate among vaccinated (adj IRR = 1.48; 95% CI: 1.09-2.00). Those with primary immunodeficiency, history of organ transplant, active tumor, and use of immunosuppressant medications were at the highest risk. For the underlying conditions and drug usage analysis, a total of 1359 and 536 unique candidate conditions and drugs available for investigation, respectively (concepts needed a minimum of 100 individuals to be considered). **Table 3** summarizes the top 10 breakthrough infections-associated condition and drug concepts. In addition to previously known conditions and drugs related to immunocompromised status (e.g., immunodeficiency disorder, valganciclovir), we found conditions and drugs related to pulmonary disease (e.g. post-inflammatory pulmonary fibrosis, albuterol) were also among those significantly contributing to the increased breakthrough infection rate. The full list of associated conditions and drug concepts is provided in **eFile 2**. We analyzed the protective effect of vaccination in the “Vax” cohort using two matched “Pre-Vax” and “Un-Vax” cohorts. When comparing the “Vax” cohort with the “Pre-Vax” cohort, the risk of COVID-19 infection in vaccinated individuals was significantly lower (adj OR = 0.12, 95% CI: 0.10– 0.13), which was also the case when stratifying by age, gender, and immunocompromised status (**eTable 1**). Similarly, we found a significant reduction in incidence rate (adj IRR = 0.42, 95% CI: 0.36-0.49) when comparing the “Vax” cohort with the “Un-Vax” cohort (**eTable 2**); Similar observations were found across age, sex and immunocompromised status subgroups.

**Table 2.** Risk factors associated with breakthrough case rate in CUIMC/NYP.

| **Risk Factors** | **IR (95% CI) per 1000 person-days** | **IRR (95% CI)<sup>1</sup>** | **p-value** | **Adjusted IRR (95% CI)<sup>2</sup>** | **p-value adj** |
| --- | --- | --- | --- | --- | --- |
| Overall | 0.16 (0.14 – 0.18) |  |  |  |  |
| Age |  |  |  |  |  |
| Age ≤ 65 | 0.16 (0.13-0.2) | Ref | Ref |  |  |
| Age > 65 | 0.15 (0.12-0.19) | 1.28 (0.916-1.78) | 0.149 |  |  |
| Race |  |  |  |  |  |
| Asian | 0.06 (0.01-0.18) | Ref | Ref |  |  |
| Black | 0.19 (0.13-0.27) | 3.25 (0.988-10.7) | 0.052 |  |  |
| White | 0.15 (0.12-0.19) | 2.9 (0.913-9.19) | 0.071 |  |  |
| Other/Unknown | 0.17 (0.13-0.21) | 2.88 (0.906-9.18) | 0.073 |  |  |
| Ethnicity |  |  |  |  |  |
| Hispanic or Latino | 0.18 (0.13-0.23) | Ref | Ref |  |  |
| Not Hispanic or Latino | 0.15 (0.12-0.18) | 0.852 (0.602-1.21) | 0.365 |  |  |
| Other/Unknown | 0.17 (0.12-0.23) | 0.913 (0.595-1.4) | 0.676 |  |  |
| Sex |  |  |  |  |  |
| Female | 0.14 (0.11-0.17) | Ref | Ref |  |  |
| Male | 0.19 (0.16-0.24) | 1.47 (1.11-1.94) | 0.008 |  |  |
| Vaccine brand |  |  |  |  |  |
| Moderna/mRNA-1273 | 0.1 (0.07-0.14) | Ref | Ref | Ref | Ref |
| Pfizer/BNT162b2 | 0.19 (0.16-0.22) | 1.65 (1.17-2.33) | 0.005 | 1.66 (1.17-2.35)<sup>3</sup> | 0.004 |
| Immune system |  |  |  |  |  |
| Not immunocompromised | 0.14 (0.11-0.17) | Ref | Ref | Ref | Ref |
| Is immunocompromised | 0.19 (0.15-0.24) | 1.49 (1.1-2) | 0.009 | 1.48 (1.09-2) | 0.011 |
| Active tumor | 0.22 (0.16-0.29) | 1.57 (1.11-2.21) | 0.010 | 1.56 (1.1-2.2) | 0.012 |
| CKD | 0.2 (0.13-0.29) | 1.35 (0.887-2.07) | 0.160 | 1.33 (0.864-2.06) | 0.194 |
| HIV | 0.21 (0.1-0.4) | 1.24 (0.628-2.44) | 0.538 | 1.25 (0.634-2.47) | 0.518 |
| On immunosuppressed therapy | 0.21 (0.16-0.28) | 1.46 (1.03-2.05) | 0.031 | 1.45 (1.03-2.04) | 0.033 |
| Primary immunodeficiency | 0.4 (0.21-0.68) | 2.55 (1.41-4.6) | 0.002 | 2.53 (1.4-4.58) | 0.002 |
| Organ transplant | 0.31 (0.15-0.57) | 1.9 (0.976-3.71) | 0.059 | 1.9 (0.977-3.71) | 0.058 |

<sup>1</sup> Adjusted for number of visits, days of previous observation, calendar month of the PCR test result;
<sup>2</sup> Adjusted for number of visits, days of previous observation, calendar month of the PCR test result and age at last vaccine dose;
<sup>3</sup> Adjusted for number of visits, days of previous observation, calendar month of the PCR test result, age at last vaccine dose and whether immune system is compromised;

**Table 3.**
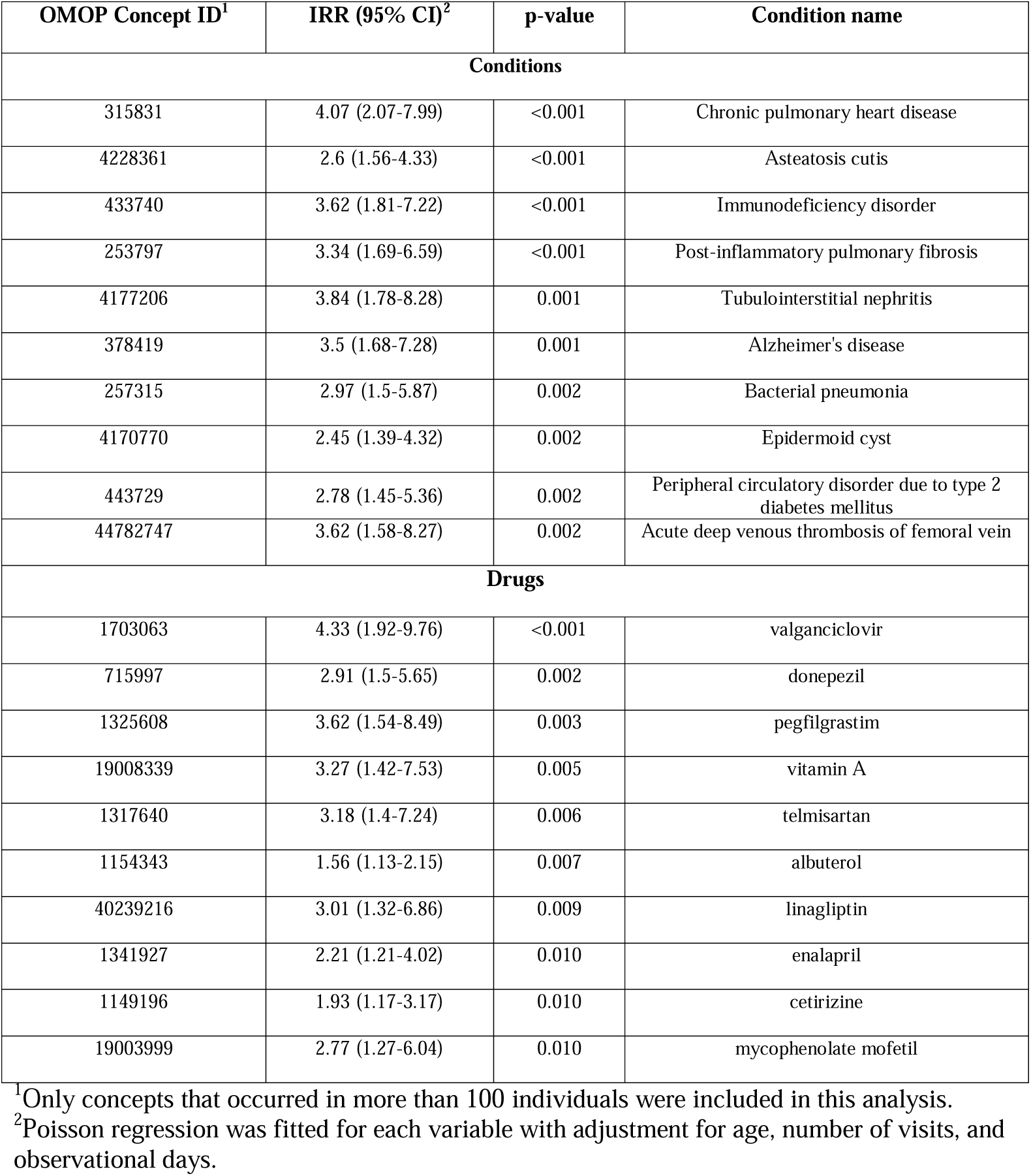
Top 10 (ranked by p-value) condition and drug concepts associated with breakthrough cases in “Vax” cohort in CUMC/NYP.

A longitudinal analysis was performed to investigate how the effectiveness of vaccines was changed over time. We calculated the cumulative incidence and incident rate at for different ranges of time to vaccination for both vaccines. Both vaccines showed an increasing incidence rate with the increased time to vaccine (**Figure 2A, 2B and eTable 3**), especially 120 days after fully vaccination. We also calculated the cumulative incidence and incident rate at different calendar month (starting from 2021-1-18; **Figure 2C, 2D** and **eTable 4**). The peaks of incidence rate observed are corresponding to the changes in the COVID variant rates in NYC, changing local mitigation measures, differences in the vaccinated population over time^17^.

**Figure 2.**
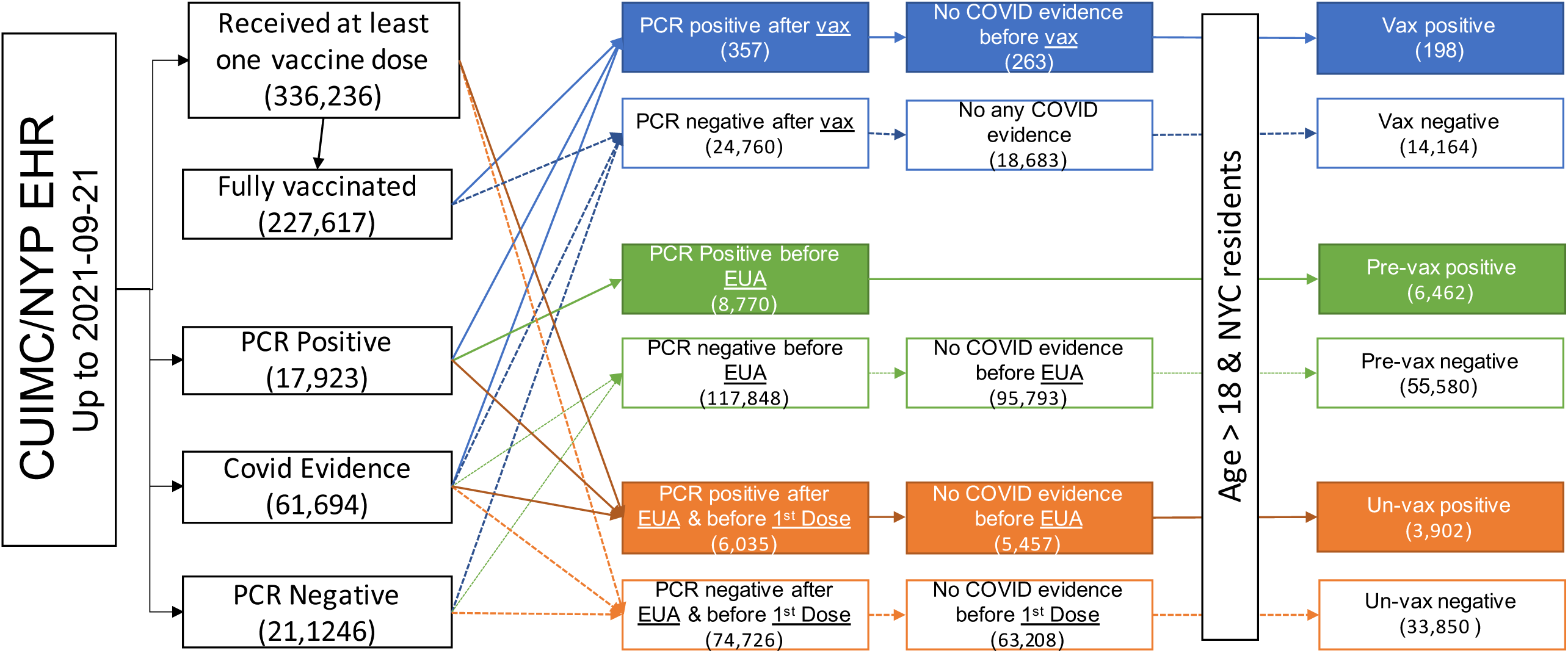
Comparison of incidence rate change along the time to vaccination and across calendar months. (A) The cumulative incidence of breakthrough cases along the time to fully vaccination; (B) The incidence rate change along the time to fully vaccination. (C) The cumulative incidence of breakthrough cases over the calendar months; (D) The incidence rate in each calendar month.

In addition to studying the effectiveness of vaccination in preventing infection, we investigated whether there were differences in the risk of experiencing severe clinical outcomes for infected individuals from the “Vax” cohort when compared to a matched “Pre-Vax” cohort (e**Table 5**) and “Un-Vax” cohort (e**Table 6**). No tracheostomies were found in breakthrough individuals. Comparing the “Pre-Vax” positive cohort to the “Vax” positive cohort, we found a significant reduction in death (adj HR = 0.20, 95% CI: 0.08-0.49). When comparing the vaccinated and unvaccinated cohorts, the hazard was significantly decreased for death (adj HR = 0.41; 95% CI: 0.17-1) and hospitalization (adj HR = 0.72; 95% CI: 0.6-0.87).

## DISCUSSION

Given the high level of missingness typically found in EHR data, it is challenging to estimate the absolute incidence rate of breakthrough infections. In our study, the incidence rate among the vaccinated cohort is estimated to be 0.16 per 1,000 person-days. This potentially overestimates the incidence rate (particularly in comparison to 0.031 in Israel’s national surveillance data^18^, ∼0.01 in the original Pfizer/BNT162b2 and Moderna/mRNA-1273 trials^19,20^) because we imposed a criterion to only include those who have at least one PCR test available, which is also called test-negative design^21^. If we remove this requirement, the incidence rate among the vaccinated cohort becomes ∼0.007 per 1,000 person-day. However, this could be underestimated because some of the SARS-CoV-2 infected individuals might have been tested elsewhere or not at all. Similarly, there is a possible proportion of patients in our unvaccinated cohort that could be vaccinated elsewhere as their vaccination records would not be captured in either our medical system or city registry, thus contributing to lower effectiveness estimations. Our longitudinal study showed a high incidence rate in a short period after full vaccination, but a chart review found many of them as asymptomatic that tested positive during admission COVID PCR screening, suggesting infection before full vaccination.

While our data support previous studies on weaker immune responses in solid fully vaccinated organ transplant recipients^22,23^, immunosuppressive therapy could be the major factor contributing to the higher incidence rate among vaccinated individuals. Despite an overall higher breakthrough infection rate in the individuals with active tumors, our sensitivity analysis found a non-statistically significant increase for individuals with history of tumors, suggesting that individuals who have fully removed their malignancy or are no longer on chemotherapy may not have a higher risk of getting infected after vaccination. An ongoing study (RECOVAC-IR study) aims to provide further guidance regarding efficacy of vaccines in kidney patients or whether other measures, like booster vaccinations, are required^24^. Our data did not find a significantly increased risk for individuals with chronic kidney disease to have a breakthrough infection, but did reveal increased risk for individuals with prior lung infection and chronic pulmonary diseases. A potential explanation is the microbiome changes within the lung that play a key role in the initiation and progression of COVID-19^25,26^. We also found individuals with Alzheimer’s disease were associated with an increased risk of infection among vaccinated individuals, which might be due to their frailty and medical vulnerability, and adhering to infection control measures such as physical distancing^27^.

While our finding reaffirmed the high protection of mRNA vaccines against COVID-19 infection, our longitudinal study found an increased incidence rate in both vaccines over the time. We found Moderna/mRNA-1273 had an overall higher effectiveness in preventing SARS-CoV-2 infections, which is consistent with a recent Mayo Clinic study up to July^28^ despite study design differences (e.g., in Mayo’s study, all vaccinated patients with or without a PCR test were included in incidence ratio calculation) and regional differences (Minnesota vs. NYC). Similarly, a recent study comparing the SARS-CoV-2 antibody response following vaccination found higher antibody titers in participants vaccinated with mRNA-1273 compared with those vaccinated with Pfizer/BNT162b2^29^. While our results indicate those vaccinated with Pfizer/BNT162b2 may need to be prioritized for a future booster shot, a recent prospective study^30^ involving 3,975 individuals up to April 2021 demonstrated no differences between vaccine effectiveness in preventing infection and found similar efficacy. Additional studies should be considered to provide further guidance on effectiveness differences between vaccine brands and booster shot prioritization.

Previous studies have shown both vaccines provide excellent protection against severe outcomes in the general population^19,20,31,32^. This protection consists of preventing infection and preventing severe outcome once individuals were infected, although it is currently not well reported whether vaccines can reduce the likelihood of developing severe outcomes once individuals are already infected. Israeli data shows that in those over the age of 65, 1,826 (32%) out of 5,686 unvaccinated infected individuals were later hospitalized due to COVID-19 compared to 451 (20.5%) out of 2,201 vaccinated infected individuals^18^. Another study found, among people aged 80 years and older, the proportion of hospitalizations is 9.14% in the vaccinated and 15.35% in the unvaccinated^33^. Our data found the hospitalization rate is ∼60% in vaccinated infected individuals and 62-78% in unvaccinated ones; this high hospitalization is likely a result of EHR data capturing sicker patients as 35-44% of the vaccinated COVID-19 negative cases were also hospitalized in our data. Additionally, our hospital system tests all patients for SARS-CoV-2 at the time of admission, meaning many were hospitalized for other reasons and could have asymptomatic or mild infections that would otherwise not have been hospitalized.

## Limitations

In addition to the aforementioned limitations of contamination in the unvaccinated cohort and inability of our data capturing individual’s entire medical history, there are other limitations. First, CUIMC/NYP is an academic medical center in NYC, which might not represent the general American population or other potential patient groups of interest. In particular, the overall population in our study are likely sicker than the general population. Second, despite adopting a test-negative design, we were still unable to confirm if negative cases were truly negative (e.g. tested positive elsewhere). Finally, we did not link our analysis with the delta variants which contributed to a surge in the caseload in New York City^14^.

## Conclusion

We performed a retrospective analysis to investigate risk factors contributing to COVID-19 breakthrough infections among vaccinated individuals. We found those who are male, immunocompromised, and with pulmonary disease are at a higher risk of COVID-19 infection after being fully vaccinated. Although both vaccines are highly effective in preventing SARS-CoV-2 infection, Moderna/mRNA-1273 was associated with a lower risk of infection than Pfizer/BNT162b2. There is a reduction of protectiveness in both vaccines over the time. Larger studies that leverage multiple medical institutions’ data are warranted to better link the PCR test results and vaccination information. Those with an OMOP instance of their data can reapply our analysis to check robustness of our results (https://github.com/WengLab-InformaticsResearch/Covid19-Breakthrough).

## Supporting information

eTables

eFigure 1

eFigure 2

eFigure 3

eFile 1

eFile 2

## Data Availability

The original EHR data used for the current study are available from the corresponding authors on reasonable requests and institutional approvals. Dr. Chunhua Weng had full access to all the data in the study and takes responsibility for the integrity of the data and the accuracy of the data analysis. The code to replicate this study in another OMOP compatible database is available in https://github.com/WengLab-InformaticsResearch/Covid19-Breakthrough

## Ethics Declaration

The study adhered to the principles set out in the Declaration of Helsinki, with informed consent from all participants. Columbia University Health Sciences IRB reviewed and approved the study (IRB AAAR3954). The analysis in this study was conducted on the de-identified data.

## Author Contribution

CL and JL equally contributed to conceptualization, data curation, formal analysis, methodology, resources, visualization, validation, and original draft. CW contributed to conceptualization and supervision. CT, AS, JRR, JHK, and JZ contributed to conceptualization and study design. KN contributed to data curation. All authors contributed to review and editing.

## Supplementary

**eFile 1**. The list of OMOP concepts used for the cohort identification, event ascertainment, and retrieval of measurements

**eFile 2**. Full list of potential underlying conditions and drug usage associated with breakthrough events in vaccinated cohorts in CUMC/NYP

**eTable 1**. Vaccine effectiveness against SARS-CoV-2 infection comparing “Vax” cohort to a matched “Pre-Vax” cohort before Dec 11th, 2020.

**eTable 2**. Vaccine effectiveness against SARS-CoV-2 infection comparing “Vax” cohort to a matched “Un-Vax” cohort after Jun 18th, 2021.

**eTable 3**. Change of Incidence rate from time to fully vaccination.

**eTable 4**. Distribution of incidence rate from January 2021 to September 2021.

**eTable 5**. Vaccine effectiveness against COVID-19 associated severe outcomes in breakthrough cohort compared to matched historical COVID-19 infection cohort.

**eTable 6**. Vaccine effectiveness against COVID-19 associated severe outcomes in the breakthrough cohort compared to a matched unvaccinated COVID-19 infection cohort.

**eFigure 1**. Diagram shows the study design in identifying risk factors for breakthrough events by comparing PCR positive cases and PCR negative cases among vaccinated individuals.

**eFigure 2**. Diagram shows the study design in assessing the vaccine effectiveness by comparing vaccinated cohort and a matched pre-vaccinated cohort

**eFigure 3**. Diagram shows the study design in assessing the vaccine effectiveness by comparing vaccinated cohort and a matched unvaccinated cohort

