## Supplementary material for "A Retrospective Analysis of COVID-19 mRNA Vaccine Breakthrough Infections – Risk Factors and Vaccine Effectiveness": eTables

**eTable 1. Vaccine effectiveness against SARS-CoV-2 infection comparing “Vax” cohort to a matched “Pre-Vax” cohort before Dec 11^th^, 2020.**

|  | **N^1^**  **(Pre-Vax/Vax)** | **Prevalence -**  **(Pre-Vax/Vax)** | **Odds Ratio (95% CI)^2^** | **Adjusted Odds Ratio (95% CI)^3^** |
| --- | --- | --- | --- | --- |
| **Overall** | 14362/14362 | 1556/198 | 0.115 (0.099-0.134) | 0.116 (0.0998-0.135) |
| **Age** |  |  |  |  |
| <= 65 | 8335/8191 | 734/111 | 0.142 (0.116-0.174) | 0.145 (0.118-0.177) |
| > 65 | 6027/6171 | 822/87 | 0.0905 (0.0724-0.113) | 0.0909 (0.0727-0.114) |
| **Sex** |  |  |  |  |
| Male | 5142/5241 | 702/88 | 0.108 (0.0862-0.135) | 0.108 (0.0865-0.136) |
| Female | 9220/9120 | 854/110 | 0.12 (0.0978-0.146) | 0.121 (0.0989-0.148) |
| **Is immune compromised** |  |  |  |  |
| True | 5287/5223 | 642/90 | 0.127 (0.101-0.159) | 0.129 (0.103-0.162) |
| False | 9075/9139 | 914/108 | 0.107 (0.0873-0.131) | 0.106 (0.0864-0.129) |

^1^ Both cohorts contained 14362 individuals in total because of 1:1 matching; Matching was based on previous visit counts, observational days, demographics, underlying immune conditions and NYC 7 days rolling average of COVID-19 cases at the PCR test date.

^2.^ Odds ratio obtained by fitting a univariate logistic regression between “Vax” cohort and a matched “Pre-Vax” cohort.

^3.^ Odds ratio obtained by fitting a logistics regression adjusted for previous number of visits and observational days.

**eTable 2. Vaccine effectiveness against SARS-CoV-2 infection comparing “Vax” cohort to a matched “Un-Vax” cohort after Jun 18^th^, 2021.**

|  | **N^1^**  **(Un-Vax/Vax)** | **Incident rate / 1000 person-days**  **(Un-Vax/Vax)** | **Incident Rate Ratio (95% CI)^2^** | **Adjusted Incident Rate Ratio (95% CI)^3^** |
| --- | --- | --- | --- | --- |
| Overall | 14362/14362 | 0.37/0.16 | 0.422 (0.362-0.493) | 0.411 (0.352-0.48) |
| Age |  |  |  |  |
| <= 65 | 9453/8191 | 0.35/0.16 | 0.47 (0.383-0.576) | 0.471 (0.384-0.579) |
| > 65 | 4909/6171 | 0.43/0.15 | 0.354 (0.279-0.449) | 0.325 (0.255-0.413) |
| **Sex** |  |  |  |  |
| Male | 5272/5241 | 0.4/0.19 | 0.489 (0.386-0.619) | 0.483 (0.381-0.612) |
| Female | 9089/9120 | 0.36/0.14 | 0.381 (0.31-0.469) | 0.368 (0.299-0.452) |
| **Is immune compromised** |  |  |  |  |
| True | 4079/5223 | 0.41/0.19 | 0.466 (0.366-0.593) | 0.432 (0.338-0.553) |
| False | 10283/9139 | 0.36/0.14 | 0.382 (0.311-0.469) | 0.375 (0.305-0.461) |

^1^ Both cohorts contained 10283 individuals in total because of 1:1 matching; Matching was based on by previous visit counts, observational days, demographics, underlying immune conditions and NYC 7 days rolling average of COVID-19 cases at the PCR test date.

^2.^ Incident rate ratio obtained by fitting a univariate Poisson regression between vaccinated cohort and a matched “Un-Vax” cohort.

^3.^ Incident rate ratio obtained by fitting a Poisson regression adjusted for previous number of visits and observational days.

^4.^ Incident rate ratio obtained by fitting a Poisson regression adjusted for previous number of visits, observational days and age at PCR test.

**eTable 3. Change of Incidence rate from time to fully vaccination.**

|  | **Pfizer/BNT162b2** | | | **Moderna/mRNA-1273** | | |
| --- | --- | --- | --- | --- | --- | --- |
| **Time to fully vaccination** | **Total person-days at risk^1^** | **Incidence** | **Incident rate / 1000 person-days** | **Total person-days at risk** | **Incidence** | **Incident rate / 1000 person-days** |
| **210-240 days** | 3074 | 6 | 1.952 | 443 | 1 | 2.257 |
| **180-210 days** | 16811 | 24 | 1.428 | 5543 | 5 | 0.902 |
| **150-180 days** | 34847 | 16 | 0.459 | 16525 | 6 | 0.363 |
| **120-150 days** | 66486 | 27 | 0.406 | 32243 | 7 | 0.217 |
| **90-120 days** | 105697 | 15 | 0.142 | 52162 | 5 | 0.096 |
| **60-90 days** | 150864 | 16 | 0.106 | 74806 | 5 | 0.067 |
| **30-60 days** | 203392 | 26 | 0.128 | 100706 | 5 | 0.050 |
| **0-30 days** | 259596 | 26 | 0.100 | 126977 | 8 | 0.063 |

^1^Incidence rate / 1000 person-days were calculated for each time interval relative to the fully vaccinated date.

|  | **Pfizer/BNT162b2** | | | **Moderna/mRNA-1273** | | | **Un-Vax** | | |
| --- | --- | --- | --- | --- | --- | --- | --- | --- | --- |
|  | **Total person-days at risk** | **Incidence** | **Incident rate / 1000 person-days^1^** | **TAR** | **Incidence** | **Incident rate / 1000 person-days** | **TAR** | **Incidence** | **Incident rate / 1000 person-days** |
| **Sep** | 15,412 | 29 | 1.88 | 6,519 | 7 | 1.07 | 40,037 | 110 | 2.75 |
| **Aug** | 73,984 | 51 | 0.69 | 33,506 | 12 | 0.36 | 126,167 | 130 | 1.03 |
| **Jul** | 114,592 | 15 | 0.13 | 54,411 | 7 | 0.13 | 179,069 | 70 | 0.39 |
| **Jun** | 147,072 | 9 | 0.06 | 71,102 | 2 | 0.03 | 233,314 | 41 | 0.18 |
| **May** | 180,498 | 11 | 0.06 | 89,184 | 8 | 0.09 | 320,118 | 86 | 0.27 |
| **Apr** | 161,808 | 20 | 0.12 | 80,769 | 2 | 0.02 | 372,957 | 140 | 0.38 |
| **Mar** | 106,281 | 18 | 0.17 | 62,219 | 3 | 0.05 | 412,916 | 111 | 0.27 |
| **Feb** | 36,536 | 3 | 0.08 | 11,695 | 1 | 0.09 | 380,115 | 43 | 0.11 |
| **Jan** | 4,584 | 0 | 0 | 0 | 0 | 0 | 190,670 | 6 | 0.03 |

**eTable 4. Distribution of incidence rate from January 2021 to September 2021.**

^1^Incidence rate / 1000 person-days were calculated each calendar month for individuals vaccinated with Pfizer/BNT162b2 or Moderna/mRNA-1273, and unvaccinated individuals.

**eTable 5. Vaccine effectiveness against COVID-19 associated severe outcomes in breakthrough cohort compared to matched historical COVID-19 infection cohort.**

|  | **Event (Pre-Vax/Vax)^1^** | **Event rate / 1000 person-days**  **(Pre-Vax/Vax)** | **Hazard Ratio (95% CI)^2^** | **Adjusted Hazard Ratio (95% CI)^3^** |
| --- | --- | --- | --- | --- |
| Hospitalization | 1071/120 | 47.58/59.2 | 1.19 (0.982-1.43) | 1.17 (0.969-1.41) |
| Mechanical Ventilation | 155/9 | 3.57/1.85 | 0.539 (0.275-1.06) | 0.518 (0.265-1.02) |
| Tracheostomy | 19/0 | 0.42/0 | 3.52e-08 (0-Inf) | 3.32e-08 (0-Inf) |
| Death | 195/5 | 4.3/1.01 | 0.235 (0.0966-0.57) | 0.2 (0.0824-0.487) |

^1^ The N of the Pre-Vax cohort will be 10 times N of the Vax because of 1:10 matching.

^2.^ Hazard ratios were obtained by fitting a univariate Cox regression between vaccinated cohort and a matched pre-vaccinated cohort.

^3.^ Hazard ratios were obtained by fitting a Cox regression adjusted for previous number of visits, observational days, age at PCR test and underlying immune conditions (binary). Individuals were censored at their last encounter or 28 days after their PCR results, whichever comes first.

**eTable 6. Vaccine effectiveness against COVID-19 associated severe outcomes in the breakthrough cohort compared to a matched unvaccinated COVID-19 infection cohort.**

|  | **Event (Un-Vax/Vax)^1^** | **Event rate / 1000 person-days**  **(Un-Vax/Vax)** | **Hazard Ratio (95% CI)^2^** | **Adjusted Hazard Ratio (95% CI)^3^** |
| --- | --- | --- | --- | --- |
| Hospitalization | 1445/120 | 93.52/59.2 | 0.726 (0.603-0.875) | 0.723 (0.6-0.872) |
| Mechanical Ventilation | 122/9 | 2.36/1.85 | 0.747 (0.38-1.47) | 0.716 (0.363-1.41) |
| Tracheostomy | 8/0 | 0.15/0 | 3.63e-08 (0-Inf) | 3.74e-08 (0-Inf) |
| Death | 115/5 | 2.19/1.01 | 0.457 (0.187-1.12) | 0.409 (0.167-1) |

^1.^ The N of the Un-Vax cohort will be 10 times N of the Vax because of 1:10 matching.

^2.^ Hazard ratios were obtained by fitting a univariate Cox regression between vaccinated cohort and a matched pre-vaccinated cohort.

^3.^ Hazard ratios were obtained by fitting a Cox regression adjusted for previous number of visits, observational days, age at PCR test and underlying immune conditions (binary). Individuals were censored at their last encounter, first dose of vaccination, whichever comes first or 28 days after their PCR results.
