## Supplementary figures and images for "A Retrospective Analysis of COVID-19 mRNA Vaccine Breakthrough Infections – Risk Factors and Vaccine Effectiveness"

### eFigure 1

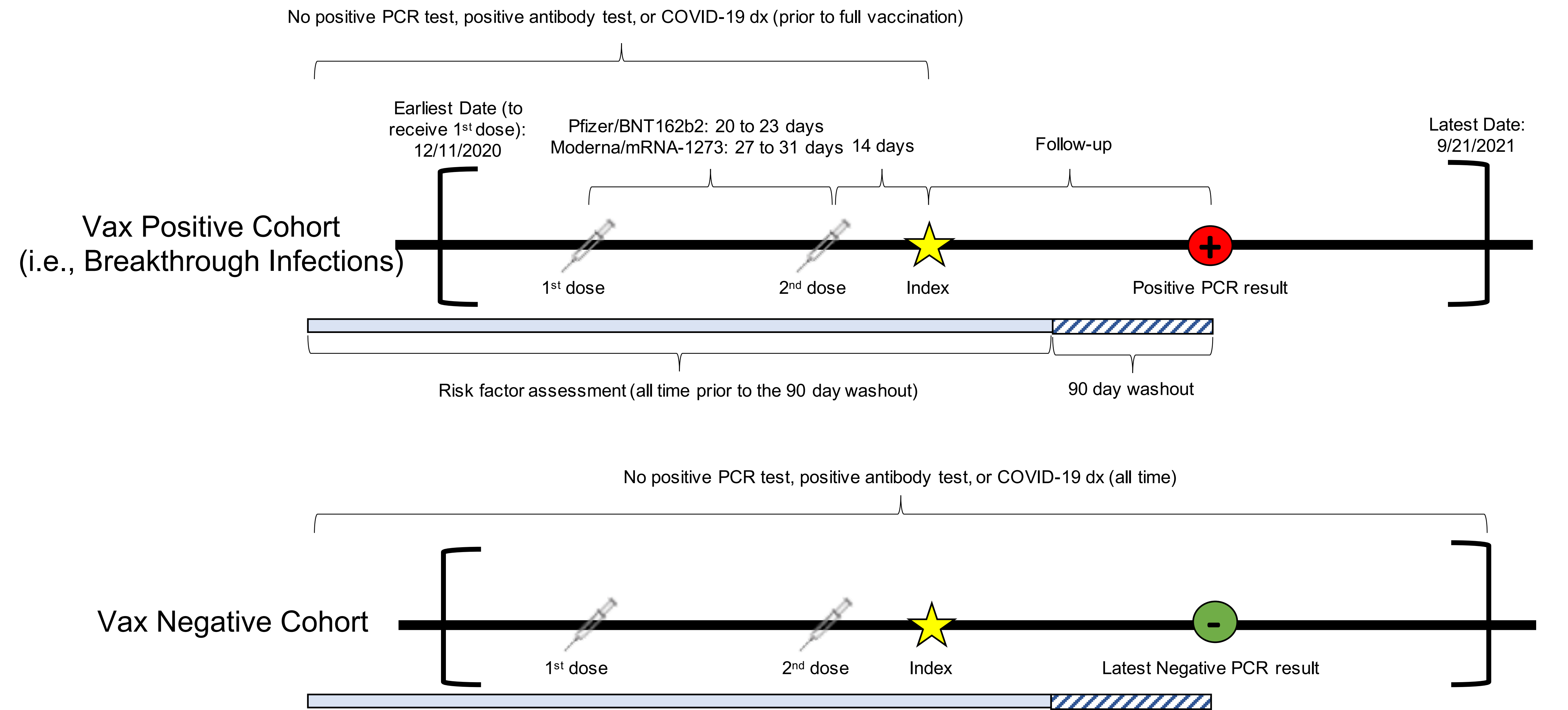

### eFigure 2

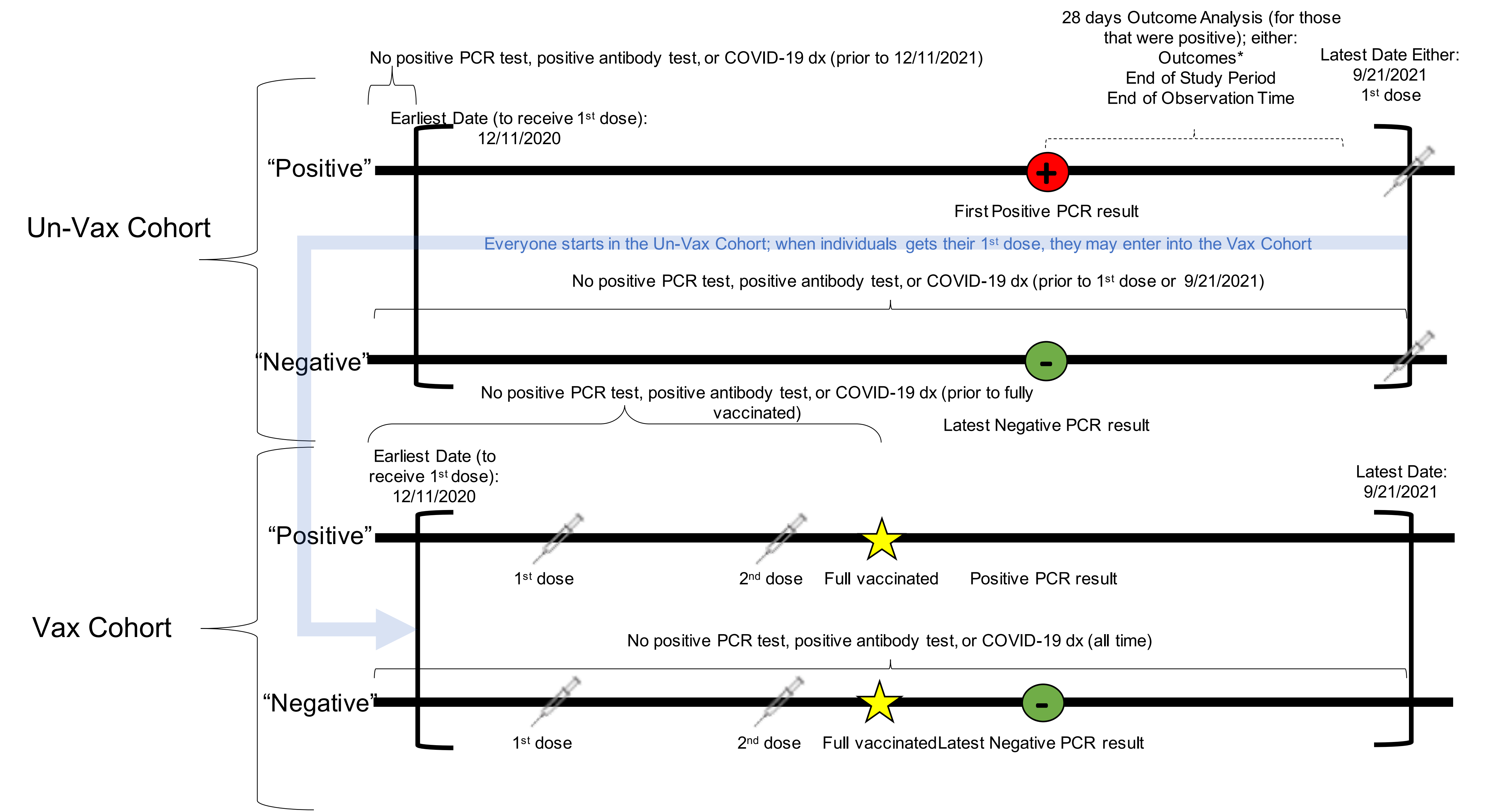


*Possible outcomes include: Ventilation, Tracheostomy, Hospitalization, Death

### eFigure 3

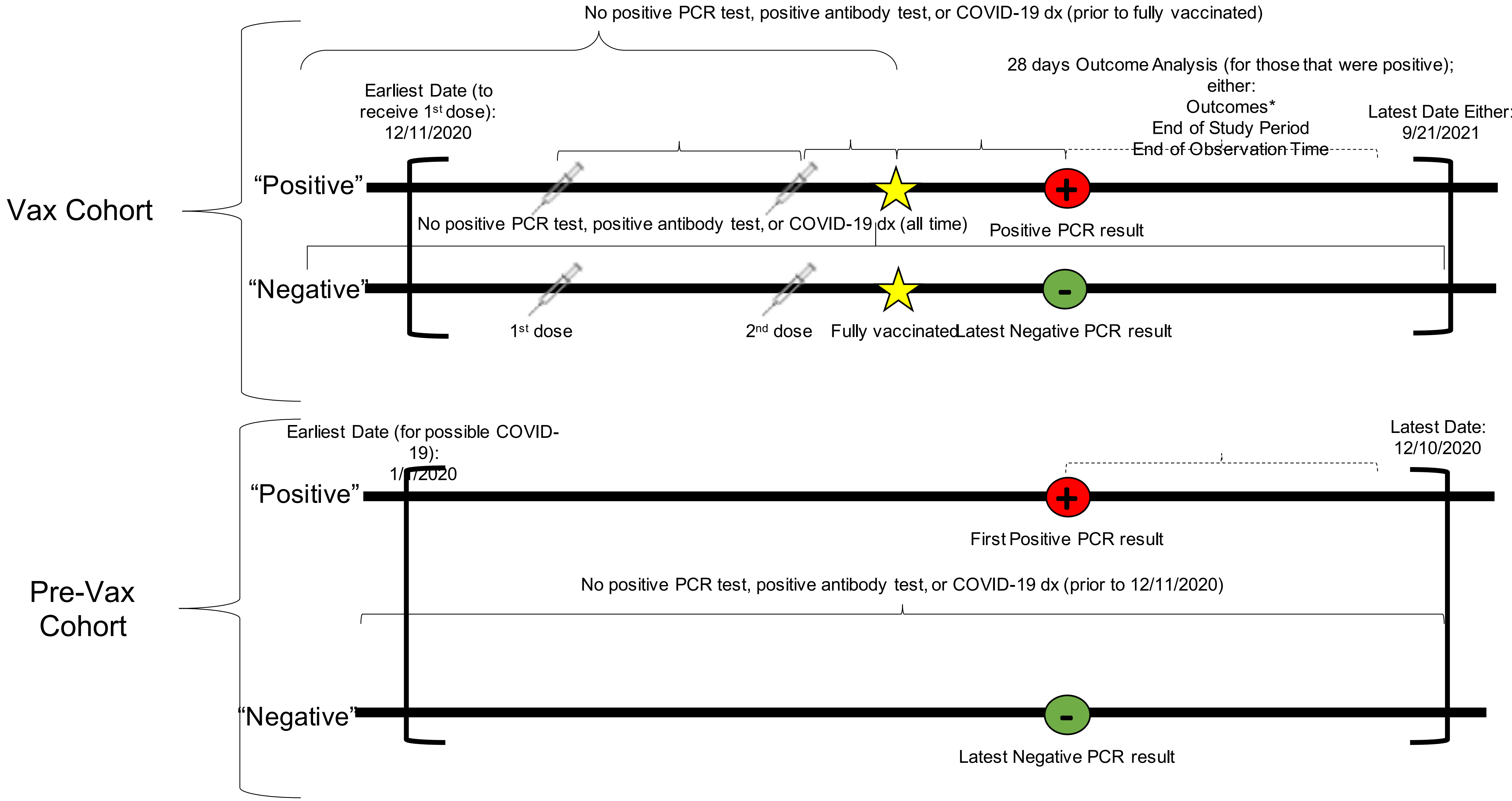


*Possible outcomes include: Ventilation, Tracheostomy, Hospitalization, Death
